# Analysis of HCM in an understudied population reveals a new mechanism of pathogenicity

**DOI:** 10.1101/2020.03.24.20037358

**Authors:** Mona Allouba, Yasmine Aguib, Roddy Walsh, Alaa Afify, Pantazis I. Theotokis, Aya Galal, Sarah Halawa, Sara Shorbagy, Ayman M. Ibrahim, Heba Sh. Kassem, Amany Ellithy, Rachel Buchan, Mohammed Hosny, Nicola Whiffin, Ahmed Elguindy, Shehab Anwer, Stuart A Cook, James S Ware, Paul J Barton, Magdi Yacoub

## Abstract

Hypertrophic Cardiomyopathy (HCM) is an inherited disease characterized by genetic and phenotypic heterogeneity. *MYH7* represents one of the main sarcomere-encoding genes associated with HCM. Missense variants in this gene cause HCM through gain-of-function actions, whereby variants produce an abnormal activated protein which incorporates into the sarcomere as a ‘poison peptide’. Here we report a frameshift variant in *MYH7*, c.5769delG, that is associated with HCM in an Egyptian cohort (3.3%) compared with ethnically-matched controls. This variant is absent from previously published large-scale Caucasian HCM cohorts. We further demonstrate strong evidence of co-segregation of c.5769delG with HCM in a large family (LOD score: 3.01). The predicted sequence of the variant *MYH7* transcript shows that the frameshift results in a premature termination codon (PTC) downstream of the last exon-exon junction of the gene that is expected to escape nonsense-mediated decay (NMD). RNA sequencing of myocardial tissue obtained from a patient with the variant during surgical myectomy confirmed the expression of the variant *MYH7* transcript. Our analysis reveals a new mechanism of pathogenicity in the understudied Egyptian population whereby distal PTC in *MYH7* may lead to the expression of an abnormal protein.

---

Hypertrophic Cardiomyopathy (HCM) represents one of the commonest inherited cardiac conditions. It is genetically and clinically heterogenous, and most commonly caused by variants in sarcomere-encoding genes, including *MYH7* which represents the second most common cause of familial HCM [1]. Missense variants in *MYH7* are believed to cause HCM through gain-of-function actions: variants produce an abnormal activated protein that incorporates into the sarcomere as a ‘poison peptide’ [2]. Haploinsufficiency in *MYH7* is not a recognised disease mechanism for HCM, and heterozygous variants that introduce premature termination codons (PTCs - i.e. nonsense, frameshift and splice variants) have not been demonstrated to be associated with this disease.

A series of 514 Egyptian patients with HCM were assessed at Aswan Heart Centre (AHC) by echocardiography and/or magnetic resonance imaging. HCM patients were sequenced on the Illumina Miseq or Nextseq platforms using the TruSight Cardio Sequencing kit, which includes 174 genes with reported roles in inherited cardiac conditions (ICC) [3]. Egyptian healthy volunteers (EHVols) were recruited to AHC for clinical assessment and genetic testing. All EHVol participants and HCM patients gave written informed consent and both studies were reviewed and approved by the institutional research ethics committee (REC)-FWA00019142 (REC codes 20151125MYFAHC_Hvol_20161027 and 20130405MYFAHC_CMR_20130330 respectively).

Rare (i.e. gnomAD filtering allele frequency (FAF) ≤4×10^−5^) protein-altering variants in validated HCM genes were selected in the analysis to allow for the identification of putative pathogenic variants. For segregation analysis, a large family of a proband carrying the *MYH7* frameshift variant was recruited to AHC for clinical assessment and genetic testing using the ICC panel. The family pedigree was created using Genial Pedigree drawing software. RNA sequencing of myocardial tissue from the proband carrying the *MYH7* frameshift variant was performed using the TruSeq Stranded Total RNA kit (Illumina). Analysis of RNA sequencing data was performed using the GATK pipeline (v4.0.8.1) and mapped reads were visualized using the Integrative Genomics Viewer (IGV).

Here we report a frameshift variant c.5769delG in *MYH7* (ENST00000355349) that is associated with HCM in a large Egyptian cohort. In a series of unrelated Egyptian patients with HCM assessed at Aswan Heart Centre, c.5769delG was found in 3.3% of patients (17 out of 514) yet was absent from locally-recruited EHVol controls (n=400, P=5.0×10^−5^), the Genome Aggregation Database (n=125,748, P=2.0×10^−41^), and the Iranome reference database (www.iranome.com, n=800). The variant was identified in one individual from the Great Middle Eastern population study [4] (n=993, P=1.2×10^−7^), but the cardiac phenotype is not known for these subjects. The variant has not been previously seen in >6000 predominantly Caucasian HCM cases [5], and has not been reported in ClinVar or the Human Gene Mutation Database.

Detailed study of a large family showed the co-segregation of c.5769delG with HCM (LOD score: 3.01), and no other rare protein-altering variants were found in other established HCM genes [5]. The proband carrying the frameshift variant presented with the disease at the age of 29 years and all affected family members carried the variant (Figure 1A).

**Figure 1:**
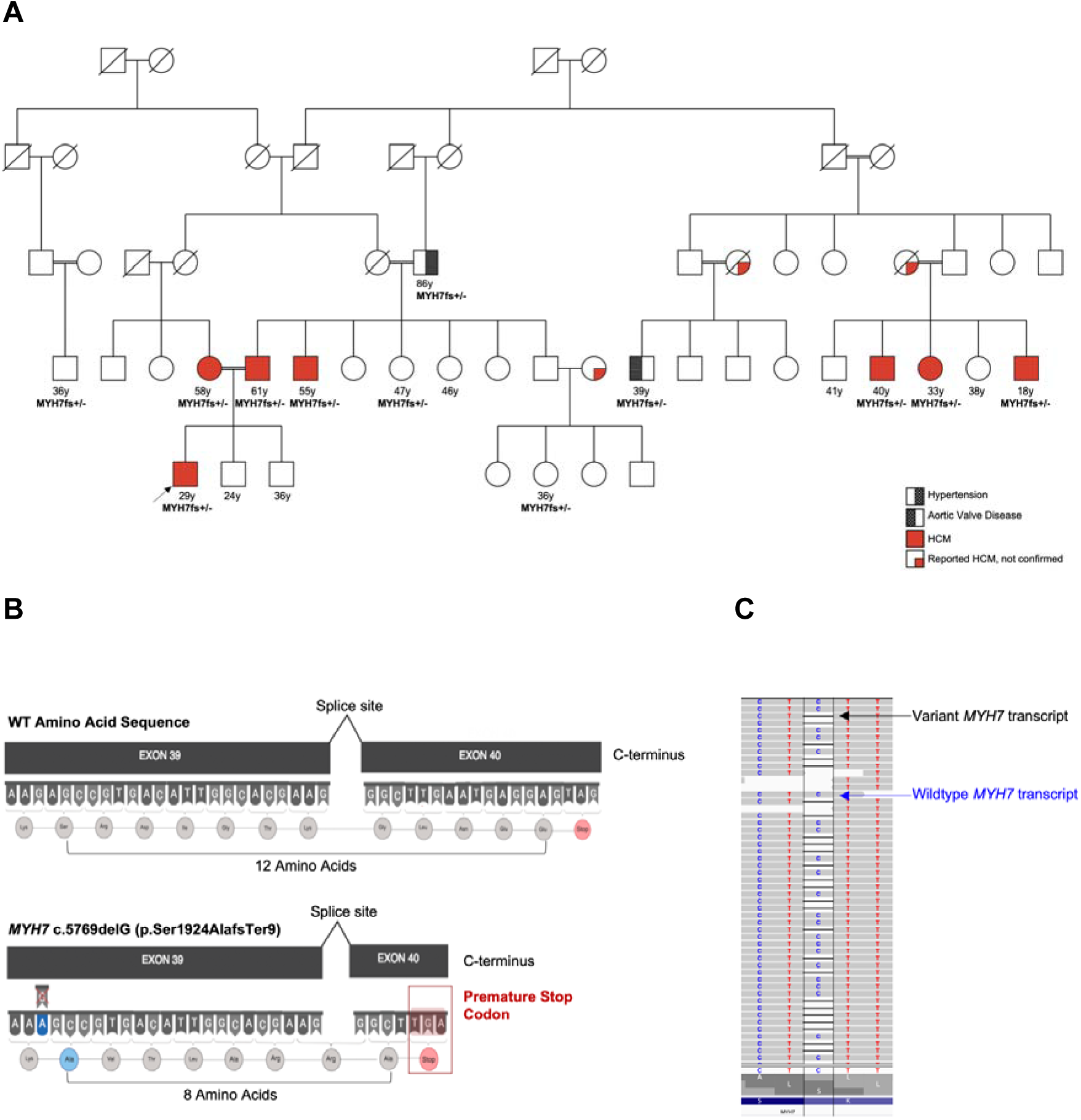
**(A) A frameshift variant in *MYH7* (c.5769delG) segregates with HCM in a large Egyptian family (LOD 3.01).** The proband is marked with an arrow. Ages of family members at diagnosis are shown. The variant segregated in all affected family members. **(B) The variant is predicted to lead to expression and translation of a truncated peptide**. The cDNA sequence and predicted resulting amino acid sequence are shown for wild type (WT) (top) and variant *MYH7* (bottom) alleles. The frameshift variant (deletion of nucleotide C in exon 39) is predicted to result in an NMD-incompetent PTC downstream of the last exon-exon junction). **(C) RNA sequencing analysis of human heart tissue from the proband confirms expression of the variant *MYH7* transcript**.

PTCs in the last exon of a gene, or in the last 50-55bp of the penultimate exon, generally escape nonsense-mediated decay (NMD), a post-transcriptional mechanism that degrades mRNAs harboring PTC [6]. The predicted sequence of the variant *MYH7* transcript shows that the PTC resides downstream of the last exon-exon junction (Figure 1B), in which case the variant transcript would be predicted to escape NMD. The translated protein sequence produces an alternate c-terminal sequence just 4 amino acids shorter than the wildtype protein. In support of this, RNA sequencing of myocardial tissue (obtained during surgical myectomy) from the proband carrying c.5769delG confirmed expression of the variant *MYH7* transcript (Figure 1C).

In *MYH7*, only missense variants have previously been proven to cause HCM. Here we demonstrate an association with an NMD-incompetent frameshift variant. While heterozygous null alleles, including NMD-competent PTC, would not be considered pathogenic for HCM in isolation (though could be potent modifiers if found *in trans* with a disease allele, or pathogenic if homozygous), distal PTC may lead to expression of an abnormal protein that may be disease-causing.

## Data Availability

Raw data from this study will be submitted to the European Genome-phenome Archive (EGA).

## Acknowledgements

This study is part of the Egyptian Collaborative Cardiac Genomics (ECCO-GEN) Project. It was supported by the Science and Technology Development Fund (STDF) government grant (Egypt), the Wellcome Trust (107469/Z/15/Z; 200990/A/16/Z), the Medical Research Council (UK), the NIHR Biomedical Research Unit in Cardiovascular Disease at Royal Brompton & Harefield NHS Foundation Trust and Imperial College London, the NIHR Imperial College Biomedical Research Centre a Health Innovation Challenge Fund award from the Wellcome Trust and Department of Health, UK (HICF-R6–373). M. A. and S.H. are funded by Al Alfi Foundation to support their PhD degrees at Imperial College London and American University in Cairo, respectively. Y.A. is supported by Fondation Leducq (11 CVD-01). N.W. is supported by a Rosetrees and Stoneygate Imperial College Research Fellowship. R.W. is supported by an Amsterdam Cardiovascular Science Fellowship.

